# Burden of Chronic Obstructive Pulmonary Disease and Its Attributable Risk Factors in China from 1990 to 2021, with Projections to 2050: An Analysis of Data from the Global Burden of Disease Study 2021

**DOI:** 10.1101/2025.03.24.25324496

**Authors:** Min Liu, Shuoshuo Wei, Xin Yang, Zhuoyuan Lu, Wanwan Zhang, Emmanuel Mensah, Lei Zha, Yun Zhou

**Affiliations:** Internal Medicine, Bengbu Medical University, Bengbu, Anhui,233000, China. Department of Pulmonary and Critical Care Medicine. The Second People’s Hospital of Wuhu, Wuhu, 241000, Anhui, China; Department of Pulmonary and Critical Care Medicine. The First Affiliated Hospital of Wannan Medical College (Yijishan Hospital of Wannan Medical College), No. 2, West Road of Zheshan, Jinghu District, Wuhu, Anhui, 241000 China; Pulmonary and Critical Care Department, The Second People’s Hospital of Wuhu, Wuhu, 241000, Anhui, China; School of Information Science & Engineering, Lanzhou University, Lanzhou, 730000, Gansu,China

**Author notes:** **Corresponding authors:** Prof. Lei Zha, Department of Pulmonary and Critical Care Medicine. The First Affiliated Hospital of Wannan Medical College (Yijishan Hospital of Wannan Medical College), No. 2, West Road of Zheshan, Jinghu District, Wuhu, Anhui, 241000 China., Prof. Yun Zhou, Department of Pulmonary and Critical Care Medicine. The Second People’s Hospital of Wuhu, Wuhu, 241000, Anhui, China.

**Keywords:** Chronic obstructive pulmonary disease, burden of disease, epidemiology, risk factors, Global Burden of Disease, China

## Abstract

**Objective:** To assess the burden of chronic obstructive pulmonary disease (COPD) and its attributable risk factors in China from 1990 to 2021 and to project the trend up to 2050 to provide a scientific basis for the development of a comprehensive COPD prevention and treatment strategy in China.

**Method:** COPD data from the Global Burden of Disease Study (GBD) 2021 were analyzed for key metrics, including incidence, prevalence, mortality, and disability-adjusted life-years (DALYs), as well as corresponding age-standardized rates (ASRs). Estimated annual percentage change (EAPC) was calculated using regression analysis, and Bayesian age-period-cohort modeling was used to project trends through 2050.

**Results:** In 2021, COPD in China reported 50,588,400 current cases of COPD, and 4,434,400 new cases, resulting in 1,285,400 million patient deaths and 23,640,300 disability-adjusted life years. The disease burden of COPD in China has generally decreased over the past 30 years. Between 1990 and 2021, the standardized incidence rate of chronic obstructive pulmonary disease (COPD) in China decreased from 271.222 per 100,000 to 215.620 per 100,000; the standardized prevalence rate declined from 2761.807 per 100,000 to 2499.374 per 100,000; the standardized mortality rate fell from 231.776 per 100,000 to 73.231 per 100,000; and the standardized DALYs (Disability-Adjusted Life Years) rate dropped from 3852.568 per 100,000 to 1227.659 per 100,000.

**Conclusion:** COPD poses a significant disease burden in China, particularly among the elderly. These findings underscore the urgency of improving diagnostic capabilities and developing better treatment strategies to address this challenging disease in the Chinese population.

## Introduction

Chronic obstructive pulmonary disease (COPD) is a heterogeneous lung condition characterized by persistent airflow limitation[1]. This limitation arises primarily from a combination of small airway diseases (e.g., obstructive bronchiolitis) and parenchymal damage (emphysema), with the relative contribution of these pathological changes varying among individuals[2]. Chronic inflammatory drives structural remodeling of the lungs, leading to narrowing of the small airways and destruction of the lung parenchyma. The resulting loss of small airway function contributes to airflow limitation and mucociliary dysfunction, hallmark features of COPD[3]. Common symptoms symptoms include progressively worsening dyspnea, cough, sputum production, wheezing, and chest tightness[4].In more severe cases, patients may also experience fatigue, weight loss, muscle wasting, and anorexia, which collectively impose a significant health and economic burden on society[5–7].

From 1990 to 2019, COPD consistently ranked high among the leading causes of death worldwide and in China. In 2019, COPD was the third leading cause of death globally, with an estimated 212.3 million prevalent cases, 3.3 million deaths, and 74.4 million disability-adjusted life years (DALYs) reported. According to the Global Burden of Disease (GBD) 2019 study, the age-standardized prevalence, mortality, and DALY rates of COPD globally in 2019 were 2,638.2 per 100,000 population (95% uncertainty interval: 2,492.2–2,796.1), 42.5 per 100,000 (95% uncertainty interval: 37.6–46.3), and 926.1 per 100,000 (95% uncertainty interval: 848.8–997.7), respectively. These figures underscore the significant public health challenges posed by COPD worldwide[8].

COPD also worsens the prognosis of other diseases, including COVID-19, cancer, mental health conditions, cardiovascular diseases, gastrointestinal disorders, and musculoskeletal diseases[9]. Beyond its severe impact on patients’ quality of life, COPD imposes a heavy economic burden globally. Between 2020 and 2050, economic losses attributed to COPD are projected to reach approximately $43.26 trillion[10]. As smoking rates rise in low- and middle-income countries and populations age in high-income countries, the prevalence of COPD is expected to increase further[11]. Projections from the Global Burden of Disease database estimate that the number of people aged ≥25 years living with COPD globally will rise by 23% from 2020 to 2050, reaching nearly 600 million by 2050, with the greatest increases anticipated among women and in low-income countries[12].

Extensive evidence links COPD to a range of risk factors, including smoking, exposure to ambient particulate matter, occupational exposure to particulates, harmful gases and fumes, household air pollution from solid fuel use, secondhand smoke exposure, ambient ozone pollution, and extreme weather conditions such as low and high temperatures[8]. Detailed definitions of these risk factors and their relative risks for COPD have been thoroughly described in previous studies[13].According to Saeid Safiri et al.[8], the primary contributors to COPD-related DALYs are smoking (46.0%), ambient particulate matter pollution (20.7%), and occupational exposure to particulates, gases, and fumes (15.6%).Systematic reviews and meta-analyses further indicates that COPD prevalence is significantly higher among smokers and former smokers compared to non-smokers, among individuals aged ≥40 years compared to those under 40, and among men compared to women[4]. Notably, COPD prevalence increases sharply with age, peaking among individuals aged ≥60 years[14].

In recent years, some studies have analyzed the status of COPD in China based on GBD data[15]. However, no studies to date have utilized the latest GBD 2021 data to assess the burden and risk factors associated with COPD in China. To address this gap, the present study leverages the most recent findings from GBD 2021 to examine trends in COPD burden and its associated risk factors in China from 1990 to 2021. Additionally, it forecasts the COPD burden from 2022 to 2050, aiming to provide a theoretical foundation for developing targeted prevention strategies and interventions for COPD..

## Methods

### Overview

We utilized data from GBD 2021 for this study, which focuses on the burden of chronic obstructive pulmonary disease (COPD) in China. The analysis covers COPD incidence, prevalence, mortality, and disability-adjusted life years (DALYs) in China from 1990 to 2021. Additionally, we conducted comparative studies based on gender and age and used a Bayesian age-period-cohort (APC) model to predict the COPD burden up to 2050.

### Data Sources

The COPD data used in this study were derived from GBD 2021 (https://vizhub.healthdata.org/gbd-results/). GBD 2021 employs the latest evidence and analytical framework from the GBD study, relying primarily on three standardized tools: the Cause of Death Ensemble model (CODEm), Spatiotemporal Gaussian Process Regression (ST-GPR), and the Disease Model–Bayesian Meta-Regression (DisMod-MR). The study comprehensively analyzes and estimates health metrics across 23 age groups, from birth to 95 years and older, for males, females, and both genders, across 204 countries and territories categorized into 21 regions and 7 super-regions. It covers health data from 1990 to 2021 and quantifies the burden of 371 diseases and injuries and 88 risk factors using metrics such as incidence, prevalence, mortality, years of life lost (YLLs), years lived with disability (YLDs), and DALYs. Age-standardized rates were calculated using the GBD standard population structure. To evaluate temporal changes, GBD 2021 presents percentage changes from 1990 to 2021, calculated as the value at the end of the time interval divided by the value at the start. All metrics’ 95% uncertainty intervals (UIs) were calculated using the mean estimates from 1,000 draws and are reported as the 2.5th and 97.5th percentiles of this distribution[16, 17].

### Case Definition

The definition of COPD included in this study follows the Global Initiative for Chronic Obstructive Lung Disease (GOLD) criteria adopted by GBD 2021: a post-bronchodilator forced expiratory volume in 1 second (FEV1)/forced vital capacity (FVC) ratio of less than 0.7. The severity of COPD was also categorized according to GOLD standards: Grade I (mild) with FEV1 ≥80% of the predicted value, Grade II (moderate) with FEV1 between 50% and 79%, Grade III (severe) with FEV1 <50%, and Grade IV (very severe) with FEV1 <30%. Other definitions of COPD include pre-bronchodilator GOLD criteria, post-bronchodilator lower limit of normal (LLN), pre-bronchodilator LLN, and guidelines from the European Respiratory Society[1, 17].

### Disease Classification

Mortality data were classified according to the International Statistical Classification of Diseases and Related Health Problems (10th Revision) (ICD-10). The disease code for COPD is J44.

### Statistical Analysis

Descriptive analyses were conducted to evaluate the COPD burden among the Chinese population aged 1 to 95 years and older. We compared the age-standardized incidence rate (per 100,000 population), prevalence rate (per 100,000 population), mortality rate (per 100,000 population), and DALY rate (per 100,000 population) across different age groups (in 5-year intervals for a total of 21 groups) and genders.

To analyze trends in age-standardized rates (ASRs) of COPD incidence, prevalence, mortality, and DALYs, we employed the Estimated Annual Percentage Change (EAPC) method. This approach involves the following regression model[18]:

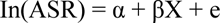

In this equation, ln(ASR)represents the natural logarithm of the age-standardized rate, Χ denotes the calendar year,α is the intercept on the y-axis, and β is the slope indicating temporal trends. Any error in the model is represented by е. EAPC is calculated as 100×[exp(β)−1], representing the annual percentage change. We analyzed trends using EAPC and its 95% CI. If the EAPC estimate and its 95% CI lower bound are both greater than 0, ASRs are considered to be increasing; if the upper bound is less than 0, ASRs are considered to be decreasing; otherwise, ASRs are deemed stable over time[18].

We also used an age-period-cohort (APC) model to assess the association between age, period, birth cohort, and COPD mortality rates[19]. The APC model is designed to unpack the contributions of age-associated biological factors, and technological as well as social factors to disease trends, extending beyond traditional epidemiological analyses[20].

The Bayesian age-period-cohort (BAPC) model was applied to predict COPD incidence, prevalence, mortality, and DALYs from 2022 to 2050. The APC model is a log-linear Poisson model that assumes multiplicative effects of age, period, and cohort, all of which follow a Poisson distribution and use a unique link function specific to this model[21]. Based on the 2021 database, we estimated prediction outcomes using 2021 COPD incidence, prevalence, mortality, and DALYs as baselines. We used an annual increase of 1% as a negative reference and an annual decrease of 1% as a positive reference[22].

Data from 1990–2021 on COPD burden in China were collected and organized using Excel 2021 software. All statistical analyses and visualizations were performed using R statistical software (version 4.4.3) and JD_GBDR (version 2.31, JD Medical Technology Co., Ltd.). A *P*-value of <0.05 was considered statistically significant.

### Risk Factors

This study included a range of risk factors associated with COPD, which have been extensively studied in previous research. These factors include smoking, ambient particulate matter pollution, occupational exposure to particulates, gases, and fumes, household air pollution caused by solid fuel use, secondhand smoke exposure, ambient ozone pollution, and extreme temperatures (both low and high)[16]. The proportion of DALYs attributable to each risk factor was analyzed. Detailed definitions of these risk factors and their relative contributions to COPD have been well-documented in prior studies[16].

## Results

### Burden of Chronic Obstructive Pulmonary Disease

In 1990, COPD was the second leading cause of death in China based on age-standardized mortality rates. By 2021, had become the third leading cause of death,accounting for 10.99% of deaths. In 2021, China reported 50.59 million prevalent cases of COPD, 4.43 million new cases, 1.29 million deaths, and 23.64 million disability-adjusted life years (DALYs). Male and female deaths accounted for 52% and 48% of the total, respectively.

Over the past three decades, the overall burden of COPD in China has shown a significant downward trend. Between 1990 and 2021, the age-standardized incidence rate (ASIR) decreased from 271.22 per 100,000 in 1990 to 215.62 per 100,000 in 2021 [EAPC: −0.843; 95% CI: −0.878 to −0.808] (**Table 1**). The age-standardized prevalence rate (ASPR) declined from 2,761.81 per 100,000 in 1990 to 2,499.37 per 100,000 in 2021 [EAPC: −0.333; 95% CI: −0.375 to −0.291] (**Table S1**). The age-standardized mortality rate (ASMR) dropped from 231.78 per 100,000 in 1990 to 73.23 per 100,000 in 2021 [EAPC: −4.250; 95% CI: −4.483 to −4.017] (**Table S2**). The age-standardized DALY rate decreased markedly from 3,852.57 per 100,000 in 1990 to 1,227.66 per 100,000 in 2021 [EAPC: −4.186; 95% CI: −4.382 to −3.990] (**Table S3**).

**Table 1.** Incidence Cases and Rates of Chronic Obstructive Pulmonary Disease (COPD) in 1990 and 2021, and the Time Trend from 1990 to 2021.

|  | Incidence(95%UI) |  |  |  | EAPC, 95%CI<br>( 1990-2021 ) |
| --- | --- | --- | --- | --- | --- |
|  | 1990 |  | 2021 |  |  |
|  | Number | rate per 100 000 | Number | rate per 100 000 |  |
| All age | 2160980.886(1992125.013,2311357.491) | 183.685(169.333,196.468) | 4434437.769(4008615.310,4860417.981) | 311.682(281.752,341.622) | 1.656(1.600,1.712) |
| age-standardi<br>zed |  | 271.222(251.664,288.622) |  | 215.620(198.000,234.903) | -0.843(-0.878,-0.808) |
| 15-19 years | 33314.176(24835.879,40015.136) | 26.301(19.607,31.591) | 14306.380(9787.752,19393.386) | 19.159(13.108,25.971) | -1.160(-1.273,-1.047) |
| 20-24 years | 47875.596(35433.222,58550.610) | 36.269(26.843,44.356) | 19113.758(13037.609,26165.056) | 26.121(17.817,35.757) | -1.276(-1.393,-1.159) |
| 25-29 years | 42136.295(30642.539,49720.889) | 38.344(27.885,45.246) | 26370.268(17287.509,34668.962) | 30.492(19.990,40.088) | -0.925(-1.022,-0.827) |
| 30-34 years | 34332.546(25575.285,40803.365) | 38.906(28.982,46.239) | 40721.808(27787.706,53141.572) | 33.612(22.936,43.863) | -0.582(-0.648,-0.516) |
| 35-39 years | 37914.085(29761.345,43243.321) | 41.509(32.583,47.344) | 40858.986(30163.733,49120.058) | 38.560(28.466,46.356) | -0.273(-0.298,-0.248) |
| 40-44 years | 87978.761(56193.697,124464.513) | 131.127(83.753,185.507) | 80836.760(54152.729,113342.933) | 88.314(59.162,123.826) | -1.426(-1.544,-1.309) |
| 45-49 years | 150358.740(111225.003,189265.627) | 291.286(215.473,366.659) | 212076.282(148430.694,290821.304) | 192.235(134.544,263.612) | -1.507(-1.583,-1.431) |
| 50-54 years | 179168.726(141761.429,210140.754) | 375.530(297.126,440.446) | 345489.392(253309.751,442322.424) | 285.861(209.591,365.982) | -0.993(-1.034,-0.952) |
| 55-59 years | 175879.798(149355.959,194532.180) | 405.542(344.383,448.550) | 392759.513(316167.821,463837.360) | 357.241(287.576,421.891) | -0.452(-0.476,-0.427) |
| 60-64 years | 275863.615(201198.502,351153.780) | 780.655(569.363,993.715) | 414650.359(310698.089,545862.356) | 567.974(425.584,747.704) | -1.089(-1.138,-1.039) |
| 65-69 years | 350317.661(272303.602,408325.106) | 1284.070(998.114,1496.694) | 729655.578(525901.722,920072.868) | 951.267(685.629,1199.519) | -1.024(-1.075,-0.973) |
| 70-74 years | 319606.954(255822.084,363905.540) | 1698.442(1359.479,1933.852) | 736483.531(546471.966,922922.541) | 1381.862(1025.344,1731.678) | -0.728(-0.787,-0.668) |
| 75-79 years | 224712.963(185049.571,250663.045) | 1974.510(1625.995,2202.528) | 574853.581(451571.691,670868.145) | 1735.725(1363.485,2025.633) | -0.507(-0.557,-0.456) |
| 80-84 years | 134991.905(109851.963,150215.170) | 2548.399(2073.803,2835.786) | 446916.235(358048.971,522564.184) | 2258.081(1809.072,2640.299) | -0.529(-0.576,-0.482) |
| 85-89 years | 52641.002(44697.888,58146.379) | 3120.659(2649.776,3447.028) | 251226.919(202535.756,289400.249) | 2637.345(2126.192,3038.083) | -0.703(-0.758,-0.649) |
| 90-94 years | 11824.712(9542.234,13663.875) | 3853.931(3110.022,4453.355) | 86624.538(68365.354,105078.031) | 2954.465(2331.707,3583.850) | -1.064(-1.130,-0.997) |
| 95+ years | 2063.351(1446.011,2541.557) | 5095.689(3571.095,6276.675) | 21493.880(14048.630,28835.852) | 3363.156(2198.195,4511.958) | -1.582(-1.673,-1.491) |
| **sex** | **Number** | **ASR\* per 100 000** | **Number** | **ASR\* per 100 000** | **EAPC** |
| Female | 1114590.105(1028463.610,1188400.257) | 269.849(251.760,286.112) | 2207589.699(1987781.195,2431561.134) | 204.657(185.881,224.539) | -0.956(-1.055,-0.856) |
| Male | 1046390.781(957619.019,1124673.577) | 274.178(252.572,293.139) | 2226848.071(2022286.573,2432575.100) | 229.215(211.290,246.901) | -0.708(-0.797,-0.618) |

Note: \*ASR, age-standardized rates;EAPC, estimated annual percentage change.

### Gender-Specific Trends in COPD Burden

The age-standardized incidence, prevalence, mortality, and DALY rates of COPD in China have consistently been higher in males than in females. However, both genders experienced a declining trend across these metrics from 1990 to 2021. During this period, the age-standardized incidence rate in males decreased from 274.18 per 100,000 in 1990 to 229.22 per 100,000 in 2021 [EAPC: −0.708; 95% CI: −0.797 to −0.618], while in females, it dropped from 269.85 per 100,000 in 1990 to 204.66 per 100,000 in 2021 [EAPC: −0.956; 95% CI: −1.055 to −0.856] (**Table 1**). Similarly, the age-standardized prevalence rate in males fell from 2,638.55 per 100,000 in 1990 to 2,479.43 per 100,000 in 2021 [EAPC: −0.335; 95% CI: −0.497 to −0.173], while in females, it decreased from 2,854.45 per 100,000 in 1990 to 2,492.11 per 100,000 in 2021 [EAPC: −0.349; 95% CI: −0.443 to −0.256] (**Table S1**).

The age-standardized mortality rate in males dropped significantly from 284.57 per 100,000 in 1990 to 105.73 per 100,000 in 2021 [EAPC: −3.571; 95% CI: −3.794 to −3.347], while in females, it declined from 199.34 per 100,000 to 52.73 per 100,000 in 2021 [EAPC: −5.002; 95% CI: −5.284 to −4.719] (**Table S2**). For DALYs, the age-standardized rate in males decreased from 4,551.32 per 100,000 in 1990 to 1,063.16 per 100,000 in 2021 [EAPC: −3.737; 95% CI: −3.919 to −3.556], while in females, it dropped from 3,358.07 per 100,000 to 963.62 per 100,000 in 2021 [EAPC: −4.671; 95% CI: −4.909 to −4.433] (**Table S3**).

### Age- and Gender-Specific Trends in COPD Burden

In 2021, the prevalence of COPD in China displayed a clear age-related trend: starting from the 15–19 age group, prevalence gradually increased, peaking in the oldest age group (≥95 years). Interestingly, while the highest number of prevalent cases was observed in the 70–74 age group, this number declined in older age group. Among individuals aged 70–74, COPD prevalence was higher in men, however, after age 74, prevalence was higher in women (**Figure 1**).

**Figure 1:**
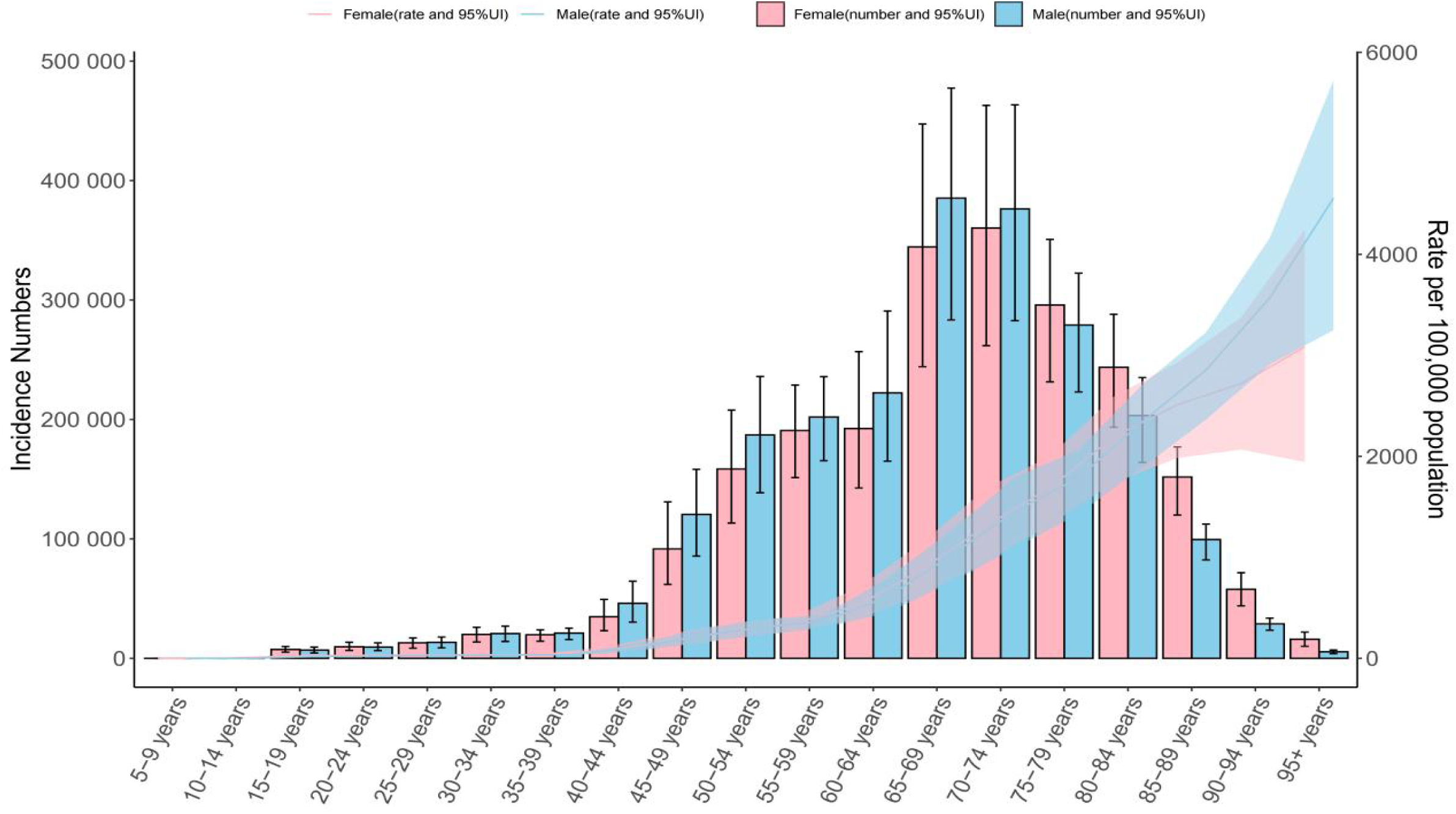
Global number of incident cases and incidence rate per 100,000 population of chronic obstructive pulmonary disease by age and sex in 2021. Note: The lines represent the number of incident cases for males and females, 95%UI.

A similar age-related pattern was observed for COPD incidence. It began to rise from the 15–19 age group, peaking in the ≥95 age group. The 70–74 age group had the highest number of new cases, but incidence rates declined beyond this age. Gender differences revealed that men aged 70–74 had higher incidence rates, while women surpassed men in incidence rates after age 74.

For COPD mortality, the highest rates were observed in the oldest age group (≥95 years) in 2021, with men consistently showing higher mortality rates than women across all age groups. In terms of absolute numbers, mortality peaked in the 80–84 age group before declining in older age groups.

Regarding DALYs, rates for men continued to rise until the 90–94 age group, after which they declined, while for women, DALYs rates continued to increase into the oldest age group (≥95 years). For all age groups under 90 years, men had higher DALY rates than women. The highest number of DALYs was recorded in the 75–79 age group, with men showing higher numbers in the 85 – 89 age group (**Figure 1, Figure S1, Figure S2, Figure S3**).

### Bayesian Age-Period-Cohort (BAPC) Model of COPD Mortality in China, 1990–2021

AFigures 2, Figure S4, and Figure S5 present the estimates from the age-period-cohort (APC) model.Between 1990 and 2021, the net drift rate for COPD mortality in China was −5.527 (95% CI: −5.854 to −5.199) for the overall population, −4.889 (95% CI: −5.312 to −4.463) for males, and −6.445 (95% CI: −6.808 to −6.081) for females.. COPD mortality generally increased with age for both men and women, reaching the highest levels in the ≥95 age group. The local drift values were −3.461 (95%CI: −4.149 to −2.769) for the overall population, −1.988 (95% CI: −3.713 to −0.232) for males, and −4.047 (95% CI: −4.563 to −3.527) for females.

**Figure 2:**
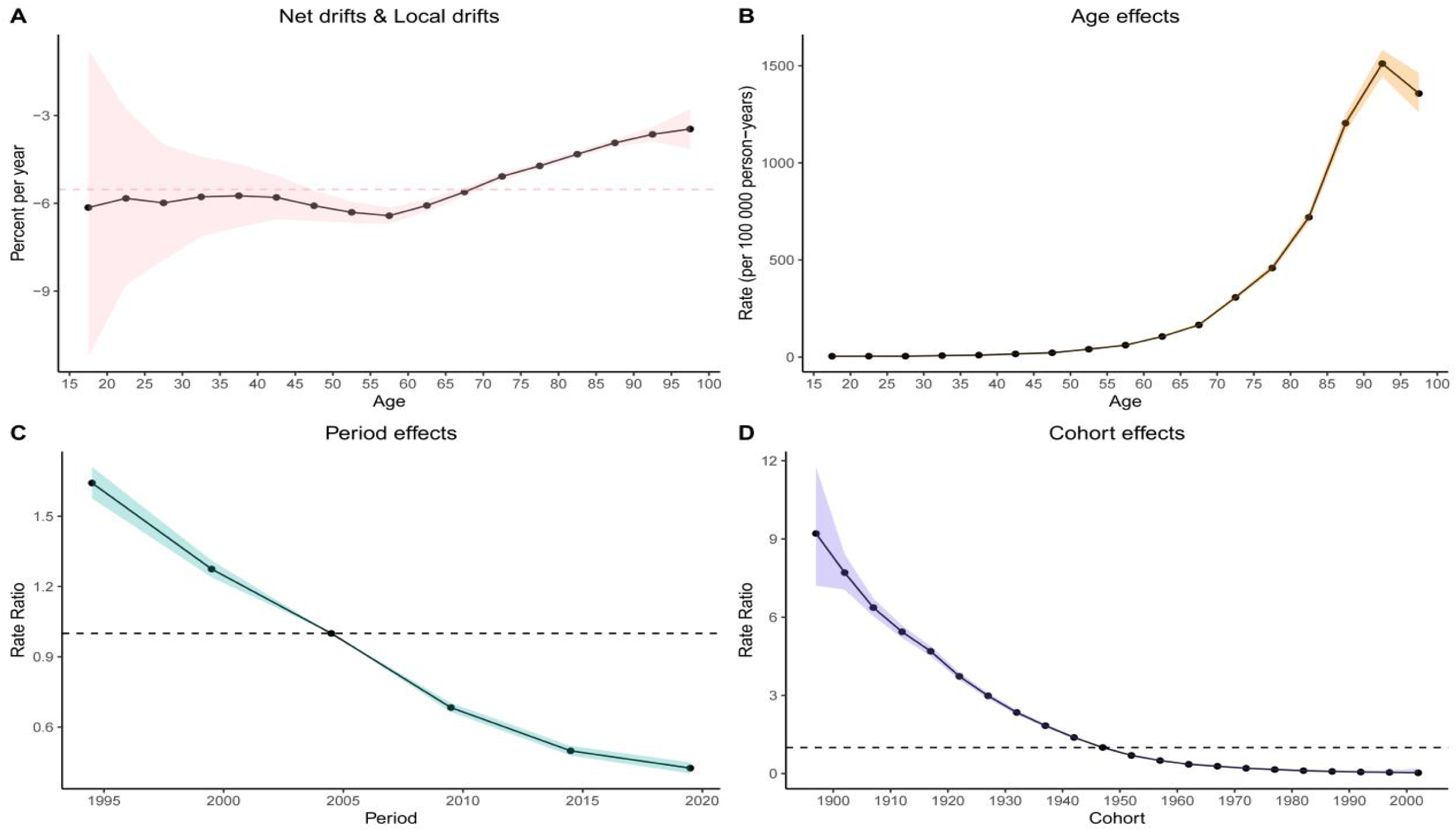
Visualization of the Bayesian Age-Period-Cohort Analysis Model for COPD Mortality in the Chinese Population from 1990 to 2021. Note: Panel A: Net drifts and Local drifts on on mortality relative risk. Panel B: Age effects on mortality relative risk;Panel C: Period effects on mortality relative risk; Panel D: Cohort effects on mortality relative risk.

**Age Effect:** After controlling for period and cohort effects, COPD mortality increased with age across the 1–95 age groups from 1990 to 2021 for both the overall population and males. However, mortality rates declined after age 95, peaking in the 90–95 age group (overall: 1,511.343 [95% CI: 1,444.325 to 1,581.472]; males: 3,161.385 [95% CI: 2,971.217 to 3,363.724]). In contrast, COPD mortality among women increased with age across all age groups, indicating a higher mortality risk among older women.

**Period Effect:** After controlling for age and cohort effects, the relative risk (RR) of COPD mortality decreased over time, with 2004.5 used as the reference value (*RR* = 1.00). This trend highlights a consistent annual decline in COPD-related mortality in China.

**Cohort Effect:** After controlling for age and period effects, the relative risk values for COPD mortality declined among both the overall population and women born between 1897 and 2002. For males, the cohort effect initially showed an increase in mortality risk for those born between 1897 and 1902, followed by a decline among cohorts born between 1903 and 2002, with the peak risk observed at *RR* = 5.557 (95% CI: 4.828 to 6.397).

### Risk Factors

The proportion of DALYs attributable to individual risk factors for COPD varied across China. Smoking (34.833%), ambient particulate matter pollution (22.161%), and indoor air pollution from solid fuels (19.500%) were the most significant contributors to COPD DALYs (**Figure 3**). The contribution of these risk factors also varied by age group.The proportion of DALYs due to smoking increased with age, peaking in the 70–74 age group before declining.DALYs attributable to ambient particulate matter pollution were highest in the 85–89 age group, though differences across other age groups were minimal.Occupational exposure to particulates, gases, and fumes accounted for an increasing proportion of COPD DALYs with age, peaking at 22.08% in the 65-69 age group **(Figure 4**). In 1990, the top-ranked contributors to COPD-related DALYs and population attributable fractions (PAFs) were indoor air pollution from solid fuels, smoking, occupational exposure to particulates, gases, and fumes, ambient particulate matter pollution, low temperatures, secondhand smoke, ambient ozone pollution, and high temperatures. Smoking surpassed indoor air pollution from solid fuels as the leading risk factor for COPD burden in 2003. By 2009, ambient particulate matter pollution became the second leading contributor, overtaking indoor air pollution from solid fuels (**Figure S8**).

**Figure 3:**
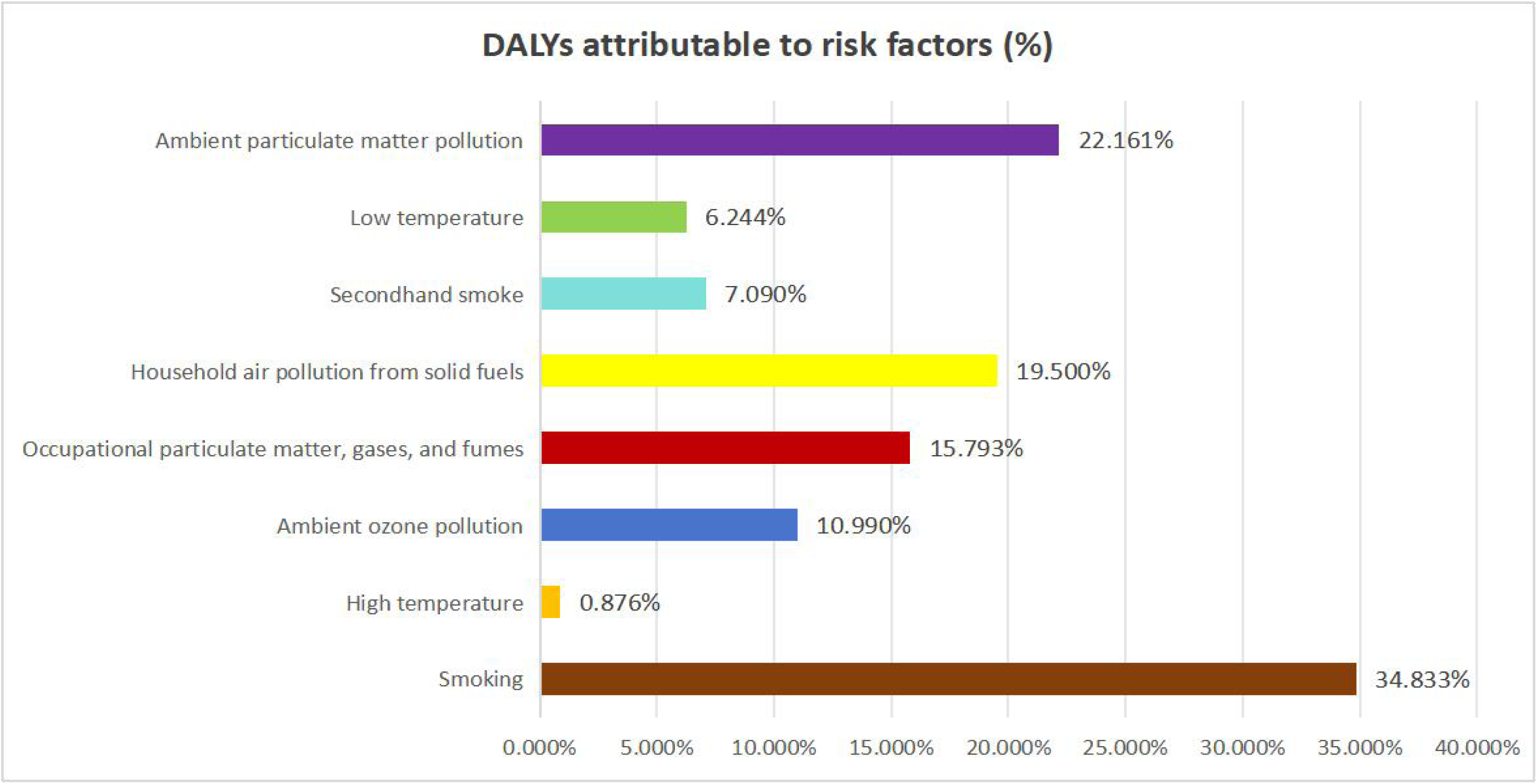
Proportion of Disability-Adjusted Life Years (DALYs) Attributable to Related Risk Factors for Chronic Obstructive Pulmonary Disease in 2021.

**Figure 4:**
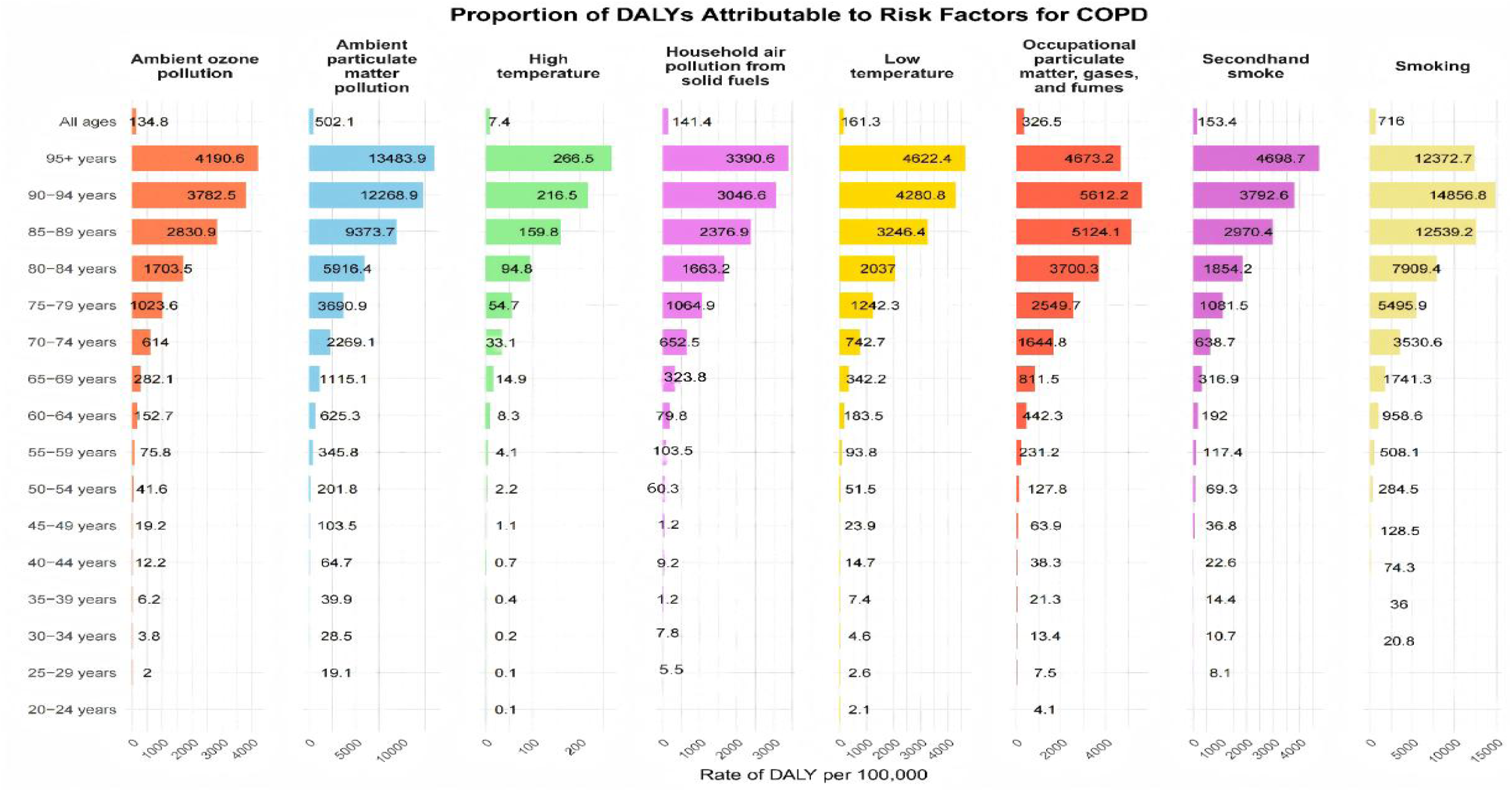
Number of Disability Cases Caused by Chronic Obstructive Pulmonary Disease Due to Risk Factors Across Different Age Groups in China, 2019.

### Projected Burden of Chronic Obstructive Pulmonary Disease in China, 2022–2050

Using the Bayesian age-period-cohort (BAPC) model, the COPD burden in China is projected to decline gradually from 2022 to 2050. By 2050, the standardized rates are expected to decrease as follows:ASIR: 193.829 per 100,000 (95% CI: 46.246 to 341.412 per 100,000); ASPR: 2,004.305 per 100,000 (95% CI: 1,871.469 to 2,137.141 per 100,000); ASMR: 50.874 per 100,000 (95% CI: −63.051 to 164.799 per 100,000); ASDR: 887.431 per 100,000 (95% CI: −683.426 to 2,458.288 per **100,000)(**Figures 5, Figure S9, Figure S10, and Figure S11).

**Figure 5:**
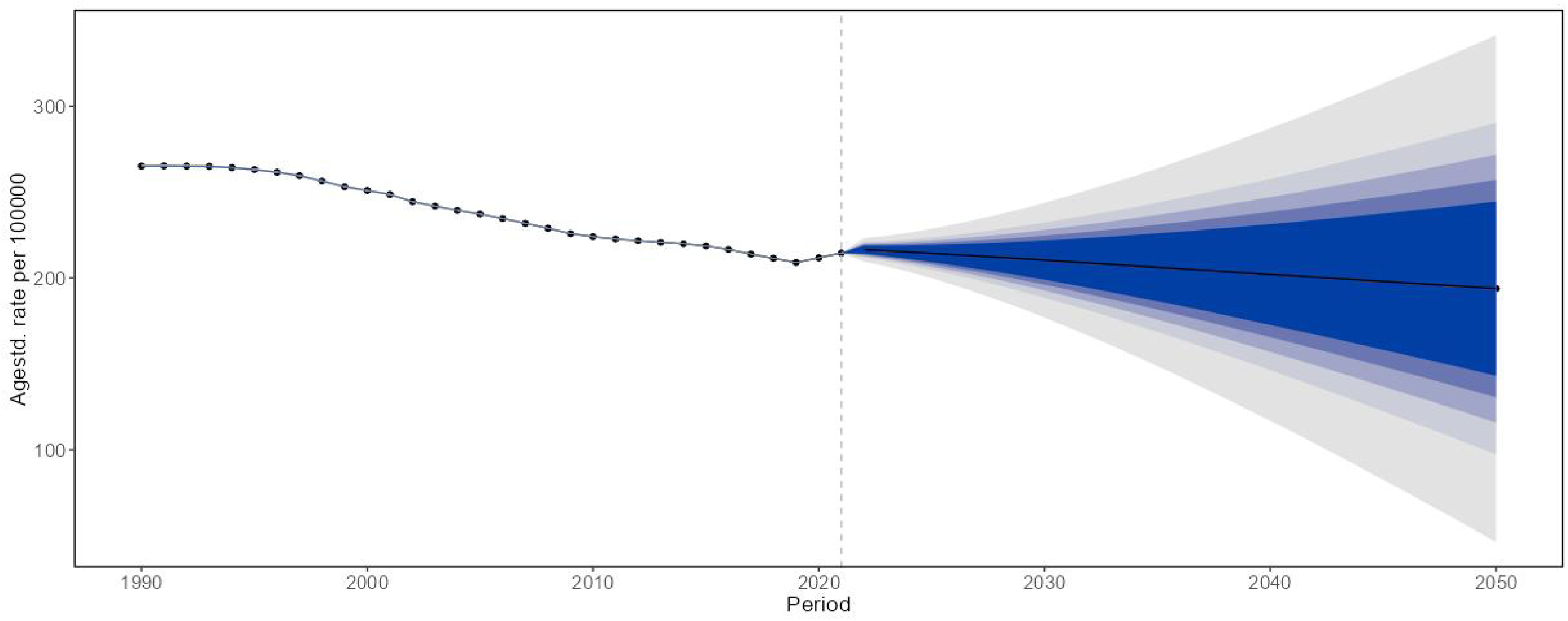
Projected Age-Standardized Incidence Rates of Chronic Obstructive Pulmonary Disease in the Overall Population of China, 2022-2050.

## Discussion

This study, based on data from the 2021 Global Burden of Disease (GBD) study, provides updated insights into COPD incidence, prevalence, mortality, and DALY counts in China from 1990 to 2021, along with age-standardized rates (ASRs). According to GBD 2021, COPD remains the fourth leading cause of death globally and the third leading cause of death in China as of 2021.

Our findings reveal that in 2021, COPD in China accounted for 4.43 million new cases, 50.59 million prevalent cases, 1.29 million deaths, and 23.64 million DALYs. While age-standardized rates for COPD incidence, prevalence, mortality, and DALYs have declined over the past 32 years, the absolute number of cases continues to rise. This trend is likely driven by factors such as population growth, aging, and increased life expectancy[23].

Over the past decade, the proportion of individuals aged ≥60 years and ≥65 years in China has risen by 5.44% (from 13.26% in 2010 to 18.70% in 2020) and 4.6% (from 8.9% to 13.5%), respectively[24].With the rapid aging of the population, China faces significant healthcare challenges[25]. By 2050, it is estimated that China will have 400 million citizens aged ≥65 years, including 150 million aged ≥80 years [26]. These demographic shifts underscore the urgent need for targeted strategies to prevent and manage COPD among middle-aged and older adults, aiming to reduce preventable cases, disabilities, and deaths..

Our results demonstrate that the burden of COPD in China increases rapidly with age, particularly among individuals aged 45 years and older, and peaks in the ≥95 age group. Physiological factors such as declining lung function, impaired lung tissue repair, and baseline inflammation in older adults may contribute to the increased risk of death[27].Additionally, COPD is associated with impaired health status[28] and multiple complications[29],which, combined with the natural comorbidities of aging, elevate mortality rates in older adults. Among COPD-related deaths, the highest number occurred in individuals aged 80–84 years, potentially due to increased vulnerability to air pollution. Previous studies have shown that air pollution has the most significant impact on COPD-related deaths among non-communicable diseases[30]. Furthermore, research indicates that the highest number of deaths attributable to air pollution occurs in individuals aged 80–84 years, mirroring the age pattern observed for COPD burden. This highlights the heightened susceptibility of older adults to the adverse effects of air pollution[30].

The study also reveals that the age-standardized prevalence, mortality, and DALY rates for COPD are higher in men than in women, reflecting differences in smoking behavior and occupational exposure to pollution. Approximately 300 million smokers in China consume 40% of the world’s cigarettes, placing Chinese men at significant risk for smoking-related disabilities and deaths. In contrast, smoking rates among Chinese women remain relatively low[31].

Our findings indicate that smoking, ambient particulate matter pollution, and occupational exposure to particulates, gases, and fumes are the largest contributors to COPD burden. These risk factors are largely preventable and manageable, suggesting that reducing the COPD burden requires more focused and coordinated efforts[32]. Smoking, in particular, is the most common risk factor for all chronic respiratory diseases[13].With nearly half of all smokers eventually developing COPD. Preventing exposure to tobacco smoke remains the most effective long-term strategy to reduce the COPD burden[33].

Studies in China have shown that non-smoking women in rural areas face a two- to three-fold higher risk of developing COPD compared to their urban counterparts due to higher exposure to biomass fuel[34].Measures such as adopting alternative clean fuels, improving kitchen ventilation, and using better stoves can significantly reduce COPD risk and improve lung function. Additionally, with the acceleration of industrialization, air pollution has become one of the greatest environmental challenges to public health in China. Evidence suggests that increases in daily concentrations of PM2.5 and PM10 are significantly associated with higher COPD incidence and reduced respiratory function[35].Strict public health measures to prevent smoking and improve air quality are critical strategies for policymakers to mitigate the COPD burden.

In 2021, the age-standardized mortality, incidence, and DALY rates for COPD in China were higher than the global averages[17].This could be attributed to the cumulative effects of risk factors such as rapid socioeconomic development, accelerated industrialization and urbanization, an aging population, and unhealthy lifestyles in recent years[36].However, the growth rates of total and new cases of COPD in China in 2021 were lower than the global averages compared to 1990[17].This suggests that China is making positive strides in preventing and managing COPD. Advances in early diagnosis and medical technologies may have contributed to the declining ASMR trend both in China and globally.

Our study found that COPD incidence, prevalence, mortality, and DALY rates in China increase significantly after age 45, peaking in the ≥95 age group. This aligns with the age effect observed in the BAPC model, which shows that both incidence and mortality rates rise with age. These findings highlight middle-aged and older adults as priority populations for COPD prevention and control. The period effect results suggest that COPD mortality risk in China has declined annually. Cohort effect results indicate that among Chinese women, mortality risk from COPD decreased among individuals born between 1897 and 2002. However, among men, mortality risk initially increased for those born between 1897 and 1902 before declining in cohorts born between 1903 and 2002. This cohort effect changes are likely influenced by disease-related factors as well as historical, social, and medical advancements, including smoking rates among men and the gradual establishment and improvement of China’s healthcare system, which has significantly reduced mortality risks for COPD and other diseases.

Projections suggest that despite the continued growth of the elderly population in China from 2022 to 2050[37], age-standardized COPD incidence and mortality rates are expected to decline. This could be attributed to a series of measures taken by China. On September 13, 2024, China incorporated COPD into its Basic Public Health Service Program, making it the third chronic disease included, following hypertension and type 2 diabetes. Through health education, smoking cessation guidance, pulmonary rehabilitation evaluation and treatment, nutritional and psychological assessments and interventions, and encouraging patient self-management, the program aims to improve screening and early diagnosis for high-risk populations, which may further optimize COPD management.

The strength of this study lies in providing the most recent and comprehensive estimates of the levels and trends related to COPD and its risk factors in China from 1990 to 2021, as well as projections for the burden of COPD up to 2050. However, this research has several limitations. Firstly, only a limited number of high-quality epidemiological databases are available for estimating the burden of COPD. Secondly, certain risk factors, such as genetic predisposition, although rare, could not be accounted for in our estimates. Thirdly, the inclusion of claims data in input records may render the evidence unreliable, as its validity heavily depends on the bias correction process. The underdiagnosis of COPD might be due to the alleviation of respiratory symptoms through activity restriction, which reduces the likelihood of patients seeking medical care. Fourthly, we were unable to access COPD data from different provinces in China, thus precluding an analysis of regional and urban-rural disparities in COPD burden. Future research should prioritize large-scale, high-quality epidemiological studies that integrate multiple data sources to more accurately assess the epidemiological characteristics of COPD in China. These limitations underscore the necessity of improving the accuracy of data collection and employing more comprehensive case definitions to provide a crucial foundation for future research and public health planning.

## Conclusion

Although the age-standardized rates (ASRs) of COPD in China have significantly declined since 1990, the continuous increase in the absolute number of cases and prevalence highlights that COPD remains a significant public health challenge in the country. Given the severe implications of an aging population, the economic burden of COPD is expected to intensify in the future.

To mitigate the burden of COPD, it is crucial to implement the following measures:

Promoting smoking cessation: Tobacco control policies and public awareness campaigns should be strengthened to reduce the prevalence of smoking, a leading risk factor for COPD.Improving environmental management: Addressing air pollution through stricter regulations and cleaner energy initiatives is vital to reduce exposure to harmful particulate matter.Enhancing lung function screening: Routine lung function tests for high-risk populations can facilitate early diagnosis and timely interventions.Developing precise health management strategies: Tailored prevention and treatment programs, especially for middle-aged and older adults, should be established to reduce preventable cases, disabilities, and deaths associated with COPD.This study underscores the importance of proactive public health measures and evidence-based strategies to alleviate the future burden of COPD in China. With continued efforts in prevention, early detection, and effective management, the impact of COPD on individual lives and the healthcare system can be significantly reduced.

## Data Availability

https://vizhub.healthdata.org/gbd-results/

## Contributors

YZ conceptualised the study. ML and SSW completed the first draft of the manuscript. XY, ZYL, WWZ and EM contributed to the study methodology. YZ and LZ was involved in the interpretation of the data. YZ, LZ and SSW provided critical comments on drafts of the manuscript. LZ was responsible for the decision to submit the manuscript. All authors participated in the review of the manuscript and read and approved the final manuscript. All authors had access to the data in the study and had final responsibility to submit for publication.

## Data sharing statement

The data from this study can be accessed openly through the GBD 2021 online database, as outlined in the Methods section.

## Acknowledgments

We acknowledge the exceptional contributions made by the collaborators of the Global Burden of Diseases, Injuries, and Risk Factors Study 2021. We sincerely appreciate the IHME institution for providing the GBD data.

## Reference

1. Celli B, Fabbri L, Criner G, Martinez FJ, Mannino D, Vogelmeier C, et al. Definition and Nomenclature of Chronic Obstructive Pulmonary Disease: Time for Its Revision. Am J Respir Crit Care Med. 2022;206(11):1317–25.

2. Hogg JC, Timens W. The pathology of chronic obstructive pulmonary disease. Annu Rev Pathol. 2009;4:435–59.

3. Barnes PJ. Inflammatory mechanisms in patients with chronic obstructive pulmonary disease. J Allergy Clin Immunol. 2016;138(1):16–27.

4. Christenson SA, Smith BM, Bafadhel M, Putcha N. Chronic obstructive pulmonary disease. Lancet. 2022;399(10342):2227–42.

5. Goertz YMJ, Looijmans M, Prins JB, Janssen DJA, Thong MSY, Peters JB, et al. Fatigue in patients with chronic obstructive pulmonary disease: protocol of the Dutch multicentre, longitudinal, observational FAntasTIGUE study. BMJ Open. 2018;8(4):e021745.

6. Schols AM, Soeters PB, Dingemans AM, Mostert R, Frantzen PJ, Wouters EF. Prevalence and characteristics of nutritional depletion in patients with stable COPD eligible for pulmonary rehabilitation. Am Rev Respir Dis. 1993;147(5):1151–6.

7. Attaway AH, Welch N, Hatipoglu U, Zein JG, Dasarathy S. Muscle loss contributes to higher morbidity and mortality in COPD: An analysis of national trends. Respirology. 2021;26(1):62–71.

8. Safiri S, Carson-Chahhoud K, Noori M, Nejadghaderi SA, Sullman MJM, Ahmadian Heris J, et al. Burden of chronic obstructive pulmonary disease and its attributable risk factors in 204 countries and territories, 1990-2019: results from the Global Burden of Disease Study 2019. BMJ. 2022;378:e069679.

9. May SM, Li JT. Burden of chronic obstructive pulmonary disease: healthcare costs and beyond. Allergy Asthma Proc. 2015;36(1):4–10.

10. Chen S, Kuhn M, Prettner K, Yu F, Yang T, Barnighausen T, et al. The global economic burden of chronic obstructive pulmonary disease for 204 countries and territories in 2020-50: a health-augmented macroeconomic modelling study. Lancet Glob Health. 2023;11(8):e1183–e93.

11. Fallahzadeh A, Sharifnejad Tehrani Y, Sheikhy A, Ghamari SH, Mohammadi E, Saeedi Moghaddam S, et al. The burden of chronic respiratory disease and attributable risk factors in North Africa and Middle East: findings from global burden of disease study (GBD) 2019. Respir Res. 2022;23(1):268.

12. Boers E, Barrett M, Su JG, Benjafield AV, Sinha S, Kaye L, et al. Global Burden of Chronic Obstructive Pulmonary Disease Through 2050. JAMA Netw Open. 2023;6(12):e2346598.

13. Collaborators GBDRF. Global burden of 87 risk factors in 204 countries and territories, 1990-2019: a systematic analysis for the Global Burden of Disease Study 2019. Lancet. 2020;396(10258):1223–49.

14. Varmaghani M, Dehghani M, Heidari E, Sharifi F, Moghaddam SS, Farzadfar F. Global prevalence of chronic obstructive pulmonary disease: systematic review and meta-analysis. East Mediterr Health J. 2019;25(1):47–57.

15. Hou SS, Shi JD, Yin X, Xu Q, Jiang F, Wang N, et al. [Disease burden of chronic obstructive pulmonary diseases in China from 1990 to 2019]. Zhonghua Liu Xing Bing Xue Za Zhi. 2022;43(10):1554–61.

16. Collaborators GBDRF. Global burden and strength of evidence for 88 risk factors in 204 countries and 811 subnational locations, 1990-2021: a systematic analysis for the Global Burden of Disease Study 2021. Lancet. 2024;403(10440):2162–203.

17. Diseases GBD, Injuries C. Global incidence, prevalence, years lived with disability (YLDs), disability-adjusted life-years (DALYs), and healthy life expectancy (HALE) for 371 diseases and injuries in 204 countries and territories and 811 subnational locations, 1990-2021: a systematic analysis for the Global Burden of Disease Study 2021. Lancet. 2024;403(10440):2133–61.

18. Chen ZF, Kong XM, Yang CH, Li XY, Guo H, Wang ZW. Global, regional, and national burden and trends of migraine among youths and young adults aged 15-39 years from 1990 to 2021: findings from the global burden of disease study 2021. J Headache Pain. 2024;25(1):131.

19. Chernyavskiy P, Little MP, Rosenberg PS. Correlated Poisson models for age-period-cohort analysis. Stat Med. 2018;37(3):405–24.

20. Bell A. Age period cohort analysis: a review of what we should and shouldn’t do. Ann Hum Biol. 2020;47(2):208–17.

21. Rosenberg PS, Check DP, Anderson WF. A web tool for age-period-cohort analysis of cancer incidence and mortality rates. Cancer Epidemiol Biomarkers Prev. 2014;23(11):2296–302.

22. Wang S, Dong Z, Wan X. Global, regional, and national burden of inflammatory bowel disease and its associated anemia, 1990 to 2019 and predictions to 2050: An analysis of the global burden of disease study 2019. Autoimmun Rev. 2024;23(3):103498.

23. Mathers CD, Loncar D. Projections of global mortality and burden of disease from 2002 to 2030. PLoS Med. 2006;3(11):e442.

24. Tu WJ, Zeng X, Liu Q. Aging tsunami coming: the main finding from China’s seventh national population census. Aging Clin Exp Res. 2022;34(5):1159–63.

25. Tu WJ, Slevin M, Zeng X. Strategies for Managing the Aging Tsunami in China: Weifang Model. J Am Geriatr Soc. 2019;67(2):403–4.

26. Fang EF, Scheibye-Knudsen M, Jahn HJ, Li J, Ling L, Guo H, et al. A research agenda for aging in China in the 21st century. Ageing Res Rev. 2015;24(Pt B):197–205.

27. Mannino DM, Davis KJ. Lung function decline and outcomes in an elderly population. Thorax. 2006;61(6):472–7.

28. Janson C, Marks G, Buist S, Gnatiuc L, Gislason T, McBurnie MA, et al. The impact of COPD on health status: findings from the BOLD study. Eur Respir J. 2013;42(6):1472–83.

29. Sin DD, Anthonisen NR, Soriano JB, Agusti AG. Mortality in COPD: Role of comorbidities. Eur Respir J. 2006;28(6):1245–57.

30. State of Global Air. 2024 [Available from: https://www.stateofglobalair.org/resources/report/state-global-air-report-2024[J].

31. Chan KH, Xiao D, Zhou M, Peto R, Chen Z. Tobacco control in China. Lancet Public Health. 2023;8(12):e1006–e15.

32. Duan RR, Hao K, Yang T. Air pollution and chronic obstructive pulmonary disease. Chronic Dis Transl Med. 2020;6(4):260–9.

33. Reitsma MB, Flor LS, Mullany EC, Gupta V, Hay SI, Gakidou E. Spatial, temporal, and demographic patterns in prevalence of smoking tobacco use and initiation among young people in 204 countries and territories, 1990-2019. Lancet Public Health. 2021;6(7):e472–e81.

34. Ran PX, Wang C, Yao WZ, Chen P, Kang J, Huang SG, et al. [The risk factors for chronic obstructive pulmonary disease in females in Chinese rural areas]. Zhonghua Nei Ke Za Zhi. 2006;45(12):974–9.

35. Sin DD, Doiron D, Agusti A, Anzueto A, Barnes PJ, Celli BR, et al. Air pollution and COPD: GOLD 2023 committee report. Eur Respir J. 2023;61(5).

36. Postma DS, Bush A, van den Berge M. Risk factors and early origins of chronic obstructive pulmonary disease. Lancet. 2015;385(9971):899–909.

37. Tatum M. China’s population peak. Lancet. 2022;399(10324):509.

